# A Genetic Risk Score for Glioblastoma Multiforme Based on Copy Number Variations

**DOI:** 10.1101/2021.01.22.21250319

**Authors:** Charmeine Ko, James P. Brody

**Affiliations:** Department of Biomedical Engineering, University of California, Irvine

## Abstract

Glioblastoma multiforme is the most common form of brain cancer. Several lines of evidence suggest that glioblastoma multiforme has a genetic basis. A genetic test that could identify people who are at high risk of developing glioblastoma multiforme could improve our understanding of this form of brain cancer.

Using the Cancer Genome Atlas (TCGA) dataset, we found common germ line DNA copy number variations in the TCGA population. We tested whether different sets of these germ line DNA copy number variations could effectively distinguish patients with glioblastoma multiforme from others in the TCGA dataset. We used a gradient boosting machine, a machine learning classification algorithm, to classify TCGA patients solely based on a set of germline DNA copy number variations.

We found that this machine learning algorithm could classify TCGA glioblastoma multiforme patients from the other TCGA patients with an area under the curve (AUC) of the receiver operating characteristic curve (AUC=0.875). Grouped into quintiles, the highest ranked quintile by the machine learning algorithm had an odds ratio of 3.78 (95% CI 3.25-4.40) higher than the average odds ratio and about 40 (95% CI 20-70) times higher than the lowest quintile.

The identification of an effective germ line genetic test to stratify risk of developing glioblastoma multiforme should lead to a better understanding of how this cancer forms. This result might ultimately lead to better treatments of glioblastoma multiforme.

## Introduction

Glioblastoma multiforme (GBM) is the most common form of brain cancer[1]. GBM was once considered a subtype of the gliomas, but evidence is accumulating that it is distinct from the other gliomas[2, 3]. Glioblastoma is an aggressive form of cancer; the median survival time is measured in months. In 2008 patients diagnosed in the US had a median survival time ranging from 31.9 months to 5.6 months, depending on their age[4]. Younger patients had longer survival times.

Several lines of evidence suggest that glioblastoma multiforme has a genetic basis. First, multiple cases of this rare disease have been reported to occur within single families[5–7]. Second, the only environmental factor associated with glioblastoma multiforme, high doses of ionizing radiation, is rare and not present for most people diagnosed with this disease[8]. Most importantly, genome wide association studies have found several SNP alleles that are present significantly more in glioblastoma multiforme patients than expected[2].

The maximum accuracy of a genetic test is a function of the heritability and prevalence of a disease[9]. The heritability of glioblastoma multiforme in a Northern European population is about 26% (95% confidence interval: 17%-35%[3]. Based on this number and the prevalence of glioblastoma multiforme in a similar population (about 2-3 per 100,000 persons), the maximum accuracy of a genetic test measured by the area under the receiver operating characteristic curve (AUC) should exceed 0.95 [9]. Tests based on SNPs do not come close to this AUC value. We set out to find how well a glioblastoma multiforme predictive DNA test based on copy number variations could perform.

## Methods

We used data from TCGA. TCGA was a project that collected germ line DNA from over 8800 patients, 499 of those patients had glioblastoma multiforme. TCGA did a thorough molecular characterization of both tumor tissue and normal tissue. In this paper, we used data only derived from peripheral normal blood collected from these patients.

The National Cancer Institute funded and oversaw TCGA. This project followed a standardized workflow called the Genome Characterization Pipeline, consisting of four major steps: Tissue Collection, Genome Characterization, Genomic Data Analysis and Data Sharing and Discovery.

Tissue source sites collected peripheral normal blood samples from patients who volunteered their samples as part of clinical trials. The Biospecimen Processing Center at Nationwide Children’s Hospital curated the samples to meet the rigorous quality standards.

The tissues were then sent to the Genome Characterization Centers: The Broad Institute specialized in DNA and performed whole genome and whole exome sequencing. The Broad Institute generated the copy number variation data. The data generated by this pipeline are publicly available in the Genomic Data Commons Data Portal for researchers all around to the world.

The copy number variation (CNV) pipeline used Affymetrix Genome-Wide Human SNP Array 6.0 data to find genomic repeats and infer the copy number of these regions[10]. It was built onto the TCGA data generated by Birdsuite; an open-source tool set created by the Broad Institute[11]. The data was processed through a circular binary segmentation analysis, which translated noisy intensity measurements into chromosomal regions of equal copy number, resulting in final output files that are segmented into genomic regions with the estimated copy number for each region [12]. These copy number values were further transformed into segment mean values also known as log base 2 ratios.

The CNV data and corresponding clinical information are stored in Google BigQuery as part of the Institute for Systems Biology’s Cancer Genomics Cloud[13]. We wrote SQL and R code to access and analyze this data.

Data analysis was performed in the statistical programming language R. We used the “bigrquery” package to download the required data subsets from the cloud storage for further manipulation to be trained in GBM models. The dataset was formatted such that every row contained one observation, i.e., a single patient, which was denoted by case-barcode, a unique identifier. Every column was defined by the genomic address of a gene segment, consisting of chromosomal location, start and end base pair positions. Each cell then contained the segment mean for the gene segment defined by its column for the patient recorded in that particular row. The value could be blank (“NA”) if the CNV had no information available. Lastly, the cancer types that the patients had been diagnosed with were recorded in another column.

We trained, tested, and validated our Gradient Boosting Machine (GBM) models using H2O, an open-source machine learning platform. The software also uses many popular machine learning algorithms, both supervised and unsupervised, such as GBM, XGBoost, Random Forest, Deep Learning, etc., and this facilitates the process of algorithm comparison to determine the most suitable algorithm. Each algorithm is equipped with extensive parameters for fine-tuning to improve model performance and handle issues such as overfitting. H2O’s GBM algorithms follow the algorithm specified by Friedman et al[14].

Germline CNVs from unmasked TCGA studies were used for GBM model training. Only the germ line genomic information from normal blood sample was included; therefore, cancers derived from hematopoietic cells, i.e., Acute Myeloid Leukemia (LAML) and Chronic Myelogenous Leukemia (LCML), were excluded. Samples originating from patients with glioblastoma multiforme were labelled GBM, while all other samples were labelled “Normal”. The models were trained with 100 trees and balanced classes, with no max depth specified, and the training was followed by ten-fold cross validation to avoid overfitting and obtain the AUC values for each model.

## Results

We have previously found that the gradient boosting algorithm performs the best on this particular dataset [15]. We used the gradient boosting algorithm exclusively in this paper.

We first sought to characterize how the performance of the classification varied with the number of features (distinct copy number variations) included. The output of the TCGA bioinformatics pipeline lists copy number variations for each person’s germline DNA sample. Each person’s copy number variation is characterized by a chromosome number, start point, end point, and a log base 2 ratio value. The log base 2 ratio value characterizes whether the particular copy number variation is a duplication, deletion, or other variation. We found that many of these copy number variations were common to many different people in the dataset.

We ranked the features (copy number variations) by the number of people in which the feature appeared, as shown in Table 1. This table, for instance, indicates that an 1154 bp copy number variation on chromosome 3 is the most common copy number variation in the dataset, appearing in 3508 of 8859 different people. We refer to this particular copy number variation as the top ranked copy number variation. Similarly, we constructed a set of the top 30, 50, 75, 100, 150, and 200 ranked copy number variations.

**Table 1.** We selected copy number variations based on the number of patients in which they appeared. These copy number variations were identified as part of the TCGA bioinformatics pipeline. This table shows the top 20, ranked by count, of the copy number variations. The location of the copy number variation is characterized by its chromosome number, start and end points (in HG38 coordinates). Then, we list the length of each copy number variation, along with the count (number of patients out of 8859 who had this copy number variation), and the mean and standard deviation of the different listed values for that copy number variation.

|  | Chromosome | Start | End | Length | Count | Mean | SD |
| --- | --- | --- | --- | --- | --- | --- | --- |
| 1 | 3 | 68697108 | 68698262 | 1154 | 3508 | -0.52 | 1.32 |
| 2 | 7 | 54318812 | 54318824 | 12 | 2956 | -0.46 | 1.86 |
| 3 | 8 | 39378084 | 39529446 | 151362 | 2830 | -0.68 | 1.49 |
| 4 | 13 | 71903417 | 71906418 | 3001 | 2815 | -1.40 | 1.83 |
| 5 | 5 | 58030200 | 58037706 | 7506 | 2707 | 1.36 | 0.86 |
| 6 | Y | 2782397 | 56872112 | 54089715 | 2664 | -1.00 | 0.28 |
| 7 | 6 | 103290101 | 103314186 | 24085 | 2601 | -0.37 | 1.00 |
| 8 | 7 | 70958264 | 70961155 | 2891 | 2523 | -1.87 | 1.55 |
| 9 | 4 | 172067997 | 172068382 | 385 | 2471 | -0.70 | 1.24 |
| 10 | 5 | 104524672 | 104524903 | 231 | 2361 | -0.13 | 1.62 |
| 11 | 3 | 193160173 | 193165114 | 4941 | 2275 | -0.25 | 1.31 |
| 12 | 2 | 146106836 | 146109366 | 2530 | 2096 | -0.62 | 1.32 |
| 13 | 1 | 109690352 | 109697556 | 7204 | 2085 | 0.04 | 1.39 |
| 14 | 4 | 114254167 | 114261019 | 6852 | 2083 | -0.57 | 1.09 |
| 15 | 9 | 23363117 | 23373486 | 10369 | 2030 | -0.84 | 1.65 |
| 16 | 7 | 154601461 | 154607903 | 6442 | 2020 | -1.20 | 1.43 |
| 17 | 20 | 80664 | 1580353 | 1499689 | 1982 | 0.01 | 0.02 |
| 18 | 5 | 46271828 | 46273400 | 1572 | 1950 | -1.46 | 1.74 |
| 19 | 6 | 149661 | 254283 | 104622 | 1938 | 0.02 | 0.06 |
| 20 | 13 | 37497899 | 37510620 | 12721 | 1918 | -0.58 | 1.40 |
| 21 | 16 | 55764310 | 55764867 | 557 | 1900 | 2.54 | 0.92 |
| 22 | 4 | 9459504 | 9477699 | 18195 | 1810 | 0.33 | 1.33 |
| 23 | 5 | 177800550 | 177805604 | 5054 | 1784 | 2.20 | 0.97 |
| 24 | 5 | 181006225 | 181363319 | 357094 | 1776 | 0.02 | 0.06 |
| 25 | 8 | 40920333 | 40920952 | 619 | 1735 | -1.90 | 0.85 |
| 26 | 12 | 9481274 | 9575696 | 94422 | 1694 | -0.32 | 1.34 |
| 27 | 1 | 152789447 | 152796224 | 6777 | 1682 | -0.46 | 1.01 |
| 28 | 8 | 645892 | 645908 | 16 | 1645 | 2.77 | 2.55 |
| 29 | 19 | 51639099 | 51644944 | 5845 | 1643 | -1.02 | 1.22 |
| 30 | 8 | 111282050 | 111283031 | 981 | 1628 | 0.23 | 1.32 |

**Table 3.** Using 5-fold cross validation, each of the 8859 people in the dataset received a score computed by the machine learning model with only the 200 most common germ line DNA copy number variations as the input. The higher the score, the more likely the person was from the glioblastoma multiforme class. Then, the 8859 people were ranked by score from lowest to highest and partitioned into five quintiles. This table presents the number of patients with and without glioblastoma multiforme in each quintile along with the odds ratio (relative to the entire group) and the 95% confidence interval for the odds ratio.

| Quintile | Normal | GBM | Total | Odds Ratio | 95% Confidence Interval |
| --- | --- | --- | --- | --- | --- |
| 1 | 1762 | 10 | 1772 | 0.10 | 0.05-0.17 |
| 2 | 1744 | 28 | 1772 | 0.27 | 0.18-0.39 |
| 3 | 1737 | 35 | 1772 | 0.34 | 0.24-0.48 |
| 4 | 1672 | 100 | 1772 | 1.00 | 0.80-1.25 |
| 5 | 1445 | 326 | 1771 | 3.78 | 3.25-4.40 |

We examined how well these overlapping sets of copy number variations could predict whether a person would have glioblastoma multiforme. Figure 1 shows how the predictive ability, quantified by the AUC, of the machine learning algorithm varies with the number of different top ranked copy number variations included in the model. We found that the classification improved with more features. This data is also displayed with the receiver operating characteristic curves shown in Figure 2.

**Figure 1.**
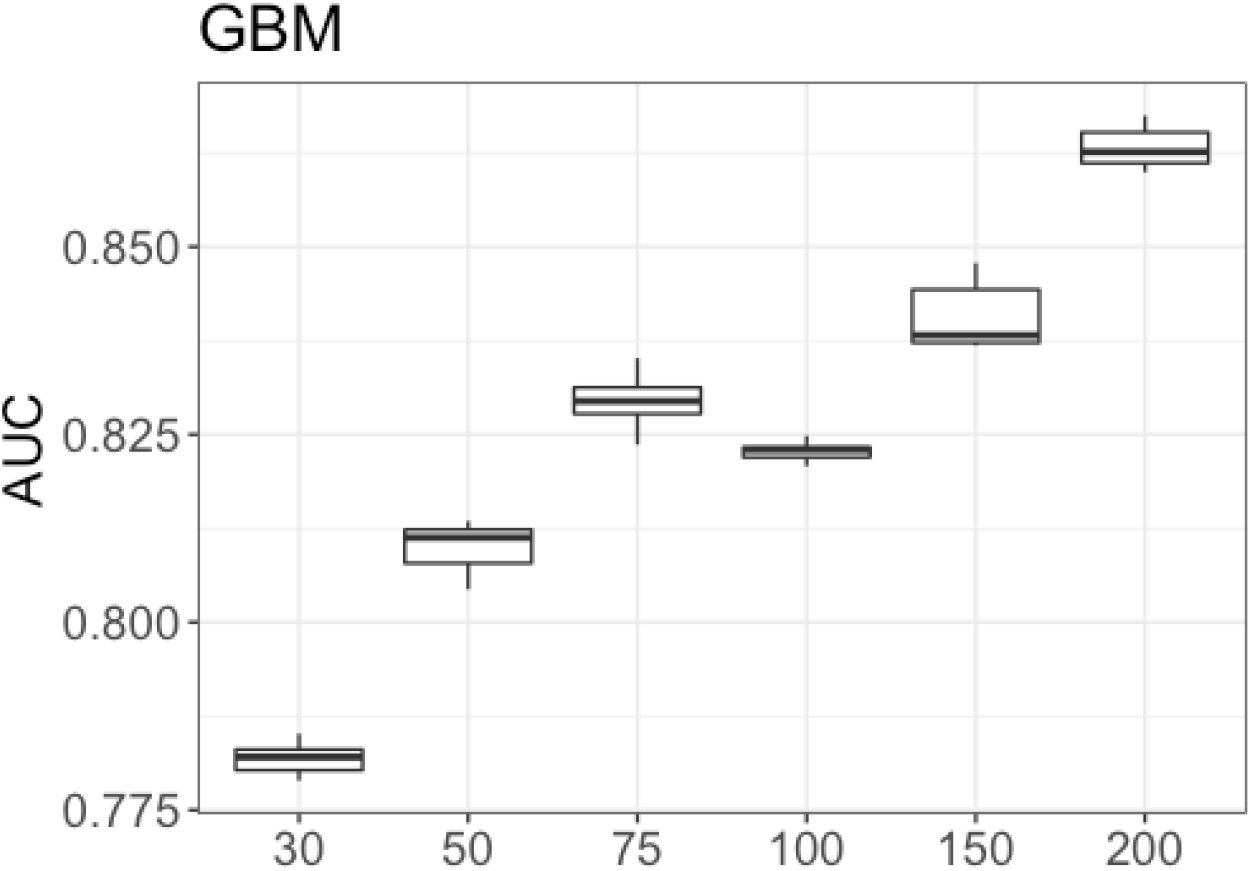
This figure quantifies how the area under the curve (AUC) of the receiver operating characteristic curve varies for six different predictive models, each using a different number of copy number variations. The x-axis indicates the number of features (distinct copy number variations) included in the model. Representative receiver operating characteristic curves for each of these models are shown in Figure 2.

**Figure 2.**
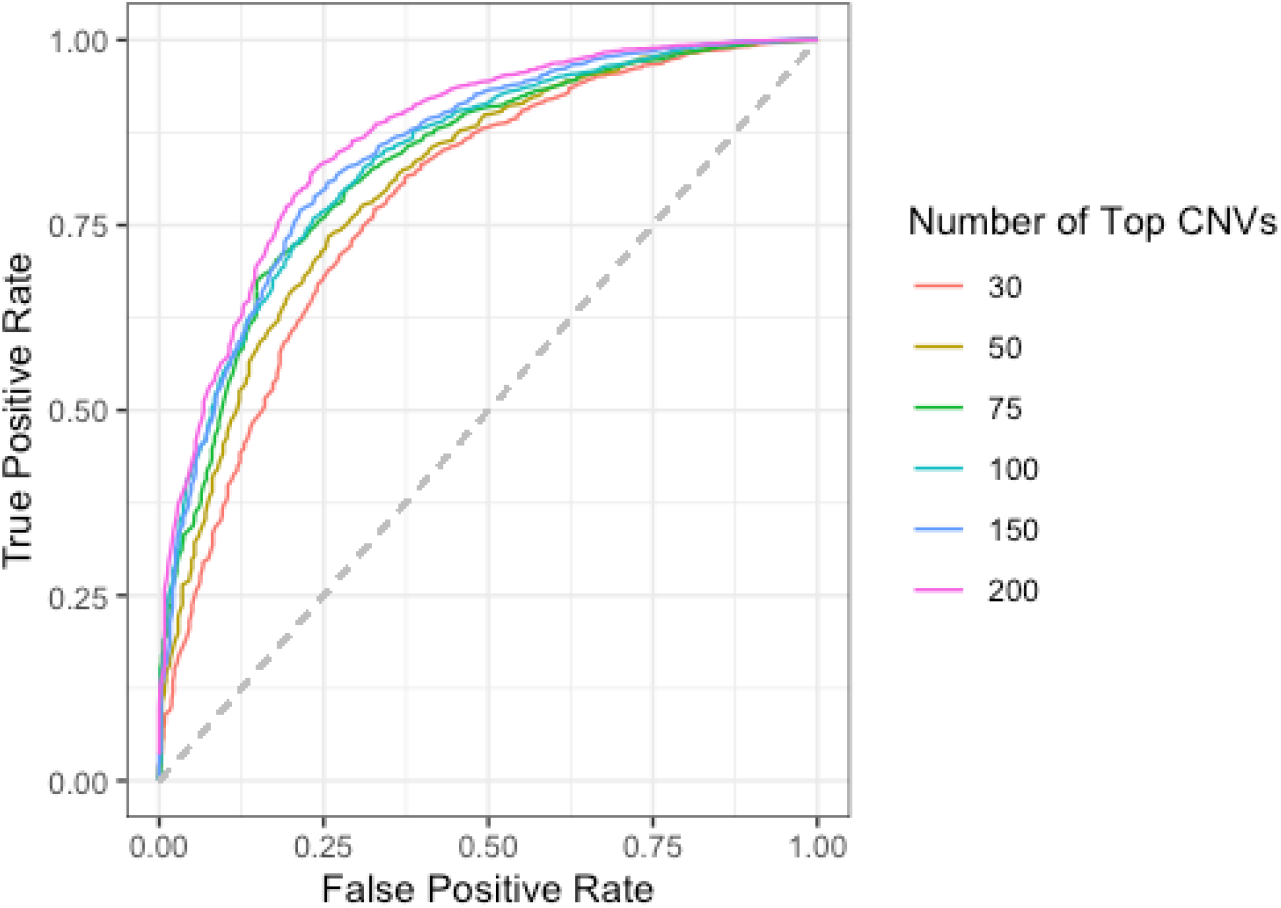
This figure presents the receiver operating characteristic curve for six different predictive models. Each model used different numbers of copy number variations to make the prediction. These receiver operating characteristic curves can be characterized by the area under the curve, AUC. The AUC for these models is shown in Figure 1.

Next, we characterized how well this classification would work as a genetic risk score[16, 17]. We used 5x cross validation to obtain genetic risk scores for each of the 8859 people in the dataset. Then, using these scores, all 8859 people were ranked by their genetic risk score and assigned a percentile. Figure 3 presents a graph of the odds ratio, relative to the entire dataset, of people in the given percentile having glioblastoma multiforme. In Figure 3, the 8859 people are binned in to 50 equal groups (2 percentile points).

**Figure 3.**
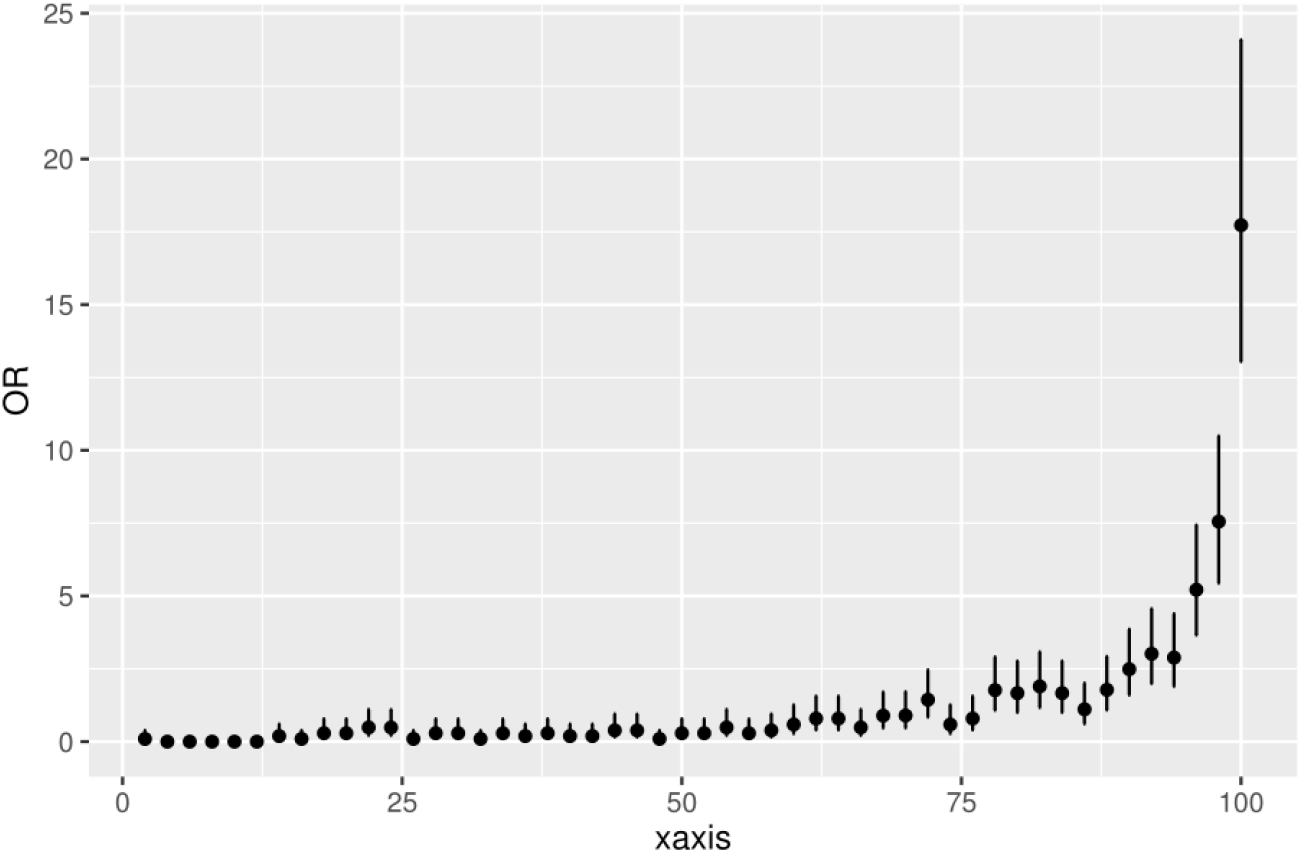
People ranked higher by the predictive model are significantly more likely to have glioblastoma multiforme. The machine learning model ranked all people in the dataset based on their likelihood of having glioblastoma multiforme. This ranking was then grouped into 50 equal partitions. This plot shows the odds ratio of each of the 50 equal partitions along with the 95% confidence intervals.

We also present this result in tabular form where the people are grouped into quintiles, five groups each consisting of 20 percentile points. This result is shown in Table 2.

## Discussion

Recent genome wide association studies have shown that glioblastoma multiforme is distinct from other forms of glioma[2]. These studies have identified a few regions of the germ line genome that are significantly different in people who develop glioblastoma multiforme. The three SNPs with the highest levels of significance are: rs10069690 at 5p15.33, rs634537 at 9p21.3 and rs2297440 at 20q13.33.

We checked whether our copy number variation data overlaps with these three SNPs. None of the three overlap with our data. The closest is the SNP rs634537, which still lies about 1.3 megabases away from the #15 copy number variation in our dataset shown in Table 1. This distance indicates the two are not related. This finding suggests that the copy number variation data is complementary to the SNP data provided by GWAS studies. The overall predictive accuracy of a germline test should increase by combining copy number variation data with SNP data.

Initial work by the TCGA project on glioblastoma multiforme focused on genome differences between normal germ line DNA and the somatic DNA found in glioblastoma tumors[18, 19]. That work identified specific mutations and complex rearrangements that most glioblastoma tumors shared. Other work on the TCGA dataset related to glioblastoma multiforme identified prognostic indicators, somatic alterations in the tumor’s DNA that could predict survival [20, 21]. In contrast, we examined only germ line DNA copy number variation to measure how well these germ line DNA alterations can predict whether a person will develop glioblastoma multiforme.

Other work on using germ line DNA copy number variation to predict development of a disease also exists. Our group used chromosomal-scale length variations, a large-scale version of copy number variation, to predict whether a person will develop ovarian cancer[15] and other forms of cancer[22] using TCGA data. We also found that the severity of response to SARS-CoV-2 infection had a small, but significant, portion that was predictable from UK Biobank chromosomal-scale length variation[23]. Another group used a GWAS-type analysis employing logistic regression with copy number variation data collected from about 1800 ovarian cancer cases and 1800 controls to demonstrate that some germ line DNA copy number variations occur more frequently in women who develop epithelial ovarian cancer than in those who do not develop that form of cancer[24].

Genetic risk scores have been developed for several other forms of cancer. One study based a genetic risk score on 53 SNPs for colorectal cancer and found that those in the highest decile of genetic risk score were 3-fold more likely to have colorectal cancer compared to the lowest decile[25]. Other studies have also found that colorectal cancers can be predicted from genetics with similar effectiveness[26, 27]. A large prostate cancer study of 1370 cases and 1239 controls found that a polygenic risk score built from 65 SNPs could predict prostate cancer with an AUC of 0.67[28]. Breast cancer can also be predicted by genetic risk scores. A recent study used a genetic risk score based on 287 SNPs in a European population and found an AUC of about 0.63. They also found that this same genetic risk score is applicable to a Chinese population, where it had an AUC of about 0.61[29]. One genetic risk score to predict breast cancer risk is commercially available and has been validated in several large cohorts with over 100,000 women. Women scoring in the top 1% of this commercially available genetic risk score have an odds ratio of about 2.0 of developing breast cancer compared to women scoring in the 40-60 percentile[30].

Our study has several limitations. We performed a reanalysis of existing data that was not collected for this purpose. It would be better to design a prospective study where samples could be collected ahead of time from a diverse, but well defined, group of people. Since we used a non-linear machine learning algorithm rather than logistic regression, we could not correct for population substructure, as is typically done in GWAS studies[31].

Our analysis is based on TCGA, a single dataset. Although TCGA was designed to be inclusive and it included a wide selection of people with different racial and ethnic backgrounds, it is not clear how well these results will generalize to any specific population. Future studies should be done to validate these findings by applying these predictions to different populations and testing how well they perform in a new population.

Finally, our control population consists of people who have been diagnosed with many different types of cancer patients, but not glioblastoma multiforme. This is an unfortunate aspect of using the TCGA dataset. It would be better to draw the control population from the general population instead of limiting it to only those who have been diagnosed with other forms of cancer.

## Conclusion

In conclusion, we found that glioblastoma multiform patients in TCGA dataset could be distinguished from other patients solely based on germline DNA copy number variation data. Germline DNA copy number variation data was able to distinguish people who developed glioblastoma from other people with an AUC of 0.875.

## Data Availability

The data in the manuscript is available from the NCI Cancer Research Data Commons.

## Acknowledgements

These results are based upon data generated by the TCGA Research Network: https://www.cancer.gov/tcga.

## Declarations

### Ethics Approval and consent to participate

Ethical approval and participant consent were collected by TCGA at the time participants enrolled. This paper is an analysis of anonymized data provided by TCGA. According to UC Irvine’s IRB, analysis of anonymized data does not constitute Human Subjects Research.

### Consent for publication

Not applicable.

### Availability of data and materials

The datasets analyzed here are available from TCGA at https://portal.gdc.cancer.gov/

### Authors’ contributions

CK and JB analyzed the TCGA data. CK and JB contributed to the manuscript. All authors read and approved the final manuscript.

